# Race and ethnic disparities in rehabilitation services and functional recovery post-stroke

**DOI:** 10.1101/2025.01.06.25320085

**Authors:** Lauri Bishop, Hannah Gardener, Scott C. Brown, Emir Veledar, Karlon H. Johnson, Erika T Marulanda-Londono, Carolina M. Gutierrez, Neva Kirk-Sanchez, Jose Romano, Tatjana Rundek

## Abstract

**Objective:** To identify race/ethnic disparities in rehabilitation services after stroke and characterize the independent associations of each of race/ethnicity and rehabilitation to functional recovery post-stroke.

**Methods:** The Transitions of Care Stroke Disparities Study (TCSD-S) is a prospective cohort study designed to reduce disparities and to optimize the transitions of care for stroke survivors throughout the state of Florida. Participant characteristics were extracted from the American Heart Association’s Get-With-The-Guidelines-Stroke dataset. Rehabilitation services, and modified Rankin Scale were recorded via follow up phone calls at 30- and 90-days after hospital discharge. Logistic regression models adjusted for potential confounders were used to determine: 1) race/ethnic differences in rehabilitation services received; 2) race/ethnic differences in functional change from discharge to 30- and 90-days, respectively; and 3) the influence of rehabilitation on functional change from discharge to 30- and 90-days.

**Results:** Of 1,083 individuals, 43% were female, 52% were Non-Hispanic White (NHW), 22% were Non-Hispanic Black (NHB), and were 22% Hispanic. Individuals who engaged in rehabilitation were more likely to show improvements [aOR=1.820, 95%CI (1.301,2.545)] at 90-days from hospital discharge. Irrespective of rehabilitation services, there were no differences in functional change between NHW and NHB individuals, yet Hispanic individuals were less likely to improve [aOR=0.647, 95%CI (0.425,0.983)] compared to NHW. Additionally, Hispanic individuals were significantly less likely to receive any rehabilitation services [aOR=0.626, 95%CI (0.442,0.886)] and were half as likely to receive outpatient services [aOR=0.543, 95%CI (0.368,0.800)] as compared to NHW.

**Conclusions:** Rehabilitation is key to functional improvement after stroke. We are making strides in health equity between NHW and NHB individuals, yet there remain disparities in functional outcomes and in rehabilitation services particularly for Hispanic individuals after stroke.

## Background

Physical activity (PA) is a critical, modifiable risk and/or protective factor of both stroke incidence and recovery post-stroke.^1–3^ Low PA (or high levels of sedentary behavior) results in a greater incidence of cardiovascular disease^2^ and further increases recurrent stroke risk.^3–5^ A key predictor of home and community PA levels after stroke is the physical ability, or physical function of the stroke survivor.^9–12^ The International Classification of Functioning, Disability and Health (ICF) highlights the interplay between physical ability, or function, activity levels and participation at home and in the community.^13^ Rehabilitation targets post-stroke motor sequelae to reduce motor impairments, increase physical ability, and thus improve physical activity in home and community environments. Further, stroke survivors consistently rank locomotor recovery as a top priority of rehabilitation.^14^ While rehabilitation efforts strive to improve physical capacity or ability, the impact of structural racism has proven a limiting factor to care access for individuals of historically disadvantaged groups.^15^

Social determinants of health (SDH) and specifically race/ethnic disparities impact stroke risk and post-stroke outcomes.^15–18^ In the United States, stroke incidence remains higher in non-Hispanic Black (NHB) and Hispanic individuals as compared to non-Hispanic Whites (NHW).^3,16–18^ In respect to post-stroke care, NHB and Hispanic individuals have been discharged home at a poorer overall functional status as compared to NHW.^19^ In order to promote health equity in functional recovery after stroke, we must first understand the current landscape of race and ethnic differences surrounding stroke rehabilitation, specifically in access and usage of post-stroke care. Therefore, the primary goal of this study was to characterize the relationship between rehabilitation and functional recovery, and further identify race/ethnic differences in functional recovery, and race/ethnic differences between types of rehabilitation services received (inpatient, outpatient rehabilitation, neither, or both).

## Methods

### Patient Population and Data Extraction

The study population was comprised of participants in the Transitions of Care Stroke Disparities-Study (TCSD-S; NIH/NIMHD R01MD012467), which has been described by prior work.^20^ TCSD-S is a prospective, multi-center observational study designed to understand and reduce disparities and to optimize the transitions of care from hospital to home or inpatient rehabilitation for stroke survivors throughout the state of Florida. Participants included stroke patients treated at 10 comprehensive stroke centers throughout the state of Florida that participate in the larger Florida Stroke Registry (a network of 181 hospitals). TCSD-S leverages data collected via forms from the American Heart Association (AHA) Get-With-The-Guidelines - Stroke (GWTG-S) during inpatient hospitalization. Structured phone interviews were completed at discharge, and at 30- and 90- days after hospital discharge in which self-report data was collected on type of stroke rehabilitation care services received (inpatient and/or outpatient rehabilitation services, or no rehabilitation) and functional status as measured by the mRS. The TCSD-S database includes 1,416 survivors of ischemic stroke or intracerebral hemorrhage who were ≥18 years of age and who were discharged either home or to an acute inpatient rehabilitation facility. Individuals with a TIA, subarachnoid hemorrhage, or other unspecified stroke diagnosis were excluded.^20^ Individuals with missing mRS at 30- and/or 90- day follow up were removed. Additionally, those who were unable to walk prior to stroke, those discharged to a skilled nursing facility or those who died during their inpatient hospital stay were removed. The study sample size was 1,083. A participant flow chart is included in **Figure 1**.

**Figure 1:**
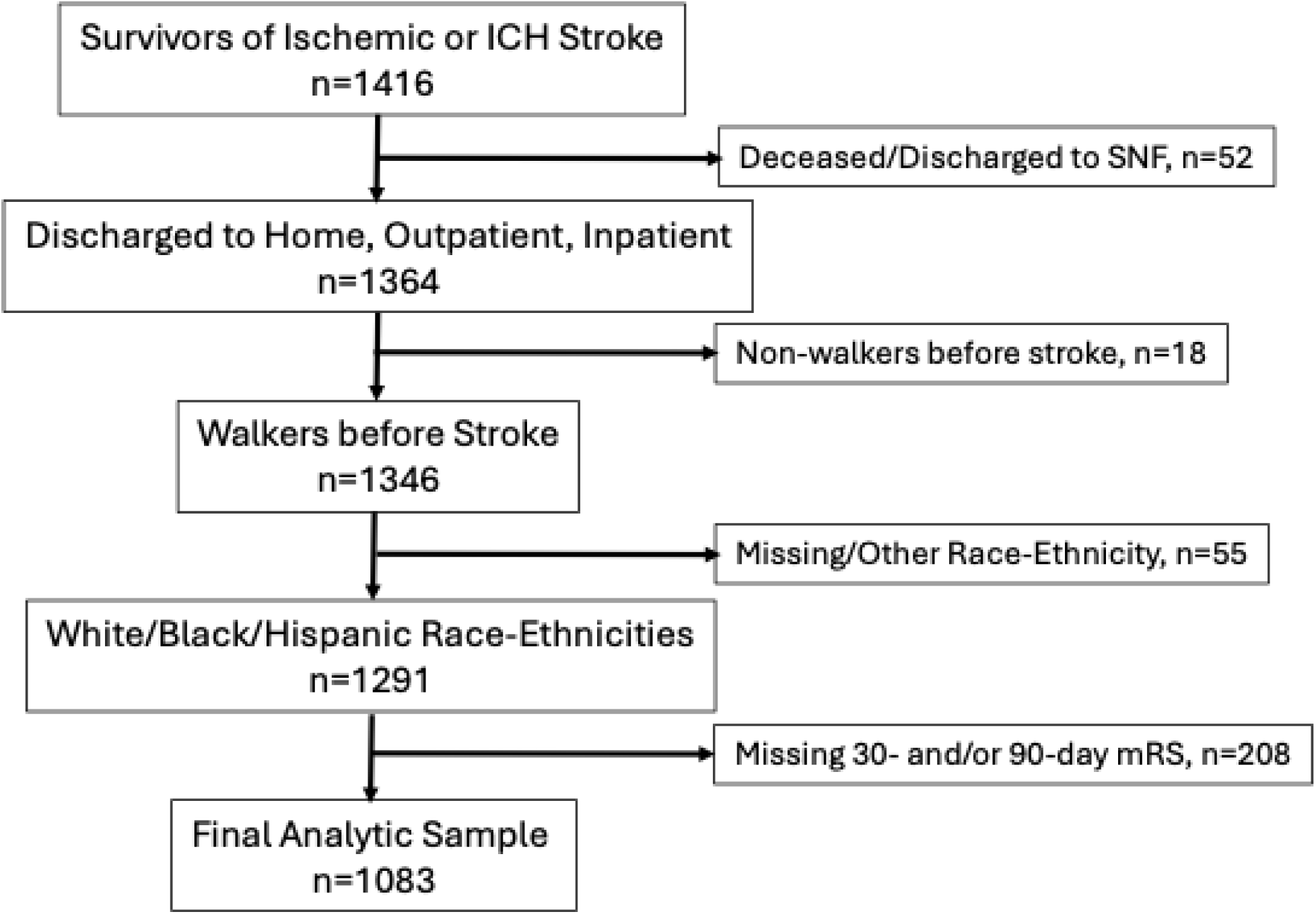
Participant flow diagram.

Race/ethnicity (NHW, NHB, Hispanic) was extracted as part of the GWTG-S database. The type of rehabilitation services in which the participant received was recorded by self-report at 30- and 90- days after discharge. Rehabilitation type was categorized by individuals who had received rehabilitation services at an inpatient rehabilitation unit (IRU), outpatient therapy (OP), both IRU and OP (Both), or received no rehabilitation services (None). Functional change (Improvement, No Change, Worse) was also recorded at 30- and 90-day follow up via self-report and was measured by a change in mRS from discharge to 30- and discharge to 90-days, respectively. Consistent with prior work^21^, ‘Improvement’ was defined as having a change in mRS from >1 at hospital discharge to an mRS of 0-1 at 30- or 90-days, respectively; OR a ≥2 point decrease in mRS from hospital discharge to 30- or 90-days, respectively. Individuals were classified as ‘Worse’ if there was an increase in mRS from hospital discharge to 30-days or from discharge to 90-days of ≥ 2, OR an increase of >1 if mRS at hospital discharge was 0-1. Individuals were categorized as ‘No Change’ if the above outlined criteria were not met.

### Statistical Analysis

Descriptive statistics (mean (SD)) are reported for continuous variables (i.e., age), and frequencies (n (%)) for categorical level data.

In order to evaluate the influence of race/ethnicity on rehabilitation participation, we first used binomial logistic regression models to determine the odds-ratios and 95% confidence intervals of going to rehabilitation (yes/no) for individuals who are NHB and those who are Hispanic compared to individuals who are NHW. We then used multinomial logistic regression models to determine the differences in type of rehabilitation services received after stroke (inpatient only, outpatient only, or both inpatient and outpatient) as compared to no therapy services by race/ethnic group (NHB/Hispanic compared to NHW). Both binomial and multinomial logistic regression models adjusted for age, sex, insurance type, stroke severity (NIHSS), walking ability at hospital discharge, social network size, who the participant lives with, ability to pay for basic necessities, and a history of prior stroke.

We then examined the influence of race/ethnicity on functional change at 30- and 90- days after discharge. Adjusting for age, sex, insurance type, stroke severity (NIHSS), walking ability at hospital discharge, social network size, who the participant lives with, ability to pay for basic necessities, and a history of prior stroke, multinomial logistic regression models were used were used to determine the odds (odds ratios and 95% confidence intervals) of improvement or worsening (compared to a reference of ‘no change’) by race/ethnic group compared to individuals who are NHW.

Due to the limited sample size and potential for confounding by indication for the analysis of rehabilitation utilization as a predictor of functional change, a propensity score analysis was conducted. Personal factors (age, sex, race, insurance type), social factors (number of individuals in support network, who the participant lives with, ability to pay for basic necessities), clinical factors (stroke severity as measured by the National Institute of Health Stroke Scale (NIHSS), walking ability at hospital discharge), and vascular factors (history of smoking, hypertension (HTN), obesity, diabetes mellitus (DM), dyslipidemia, atrial fibrillation/flutter, coronary artery disease (CAD), peripheral vascular disease (PVD), prior stroke, and depression) were used to create a propensity score that included potential covariates or influential factors on rehabilitation participation (outcome of the propensity score derivation model). We then used the propensity score as a covariate in a multinomial logistic regression model to estimate the association between rehabilitation participation (primary exposure, yes vs no) with improvement or worsening (as compared to a reference of ‘no change’) at 30- and 90-days after stroke. All statistical analyses were completed using SPSS (Released 2024. IBM SPSS Statistics for Mac, Version 29.0.2.0 Armonk, NY: IBM Corp).

## Results

The mean (SD) age of participants was 64 (14) years. Of the sample, 43% (n=482) were female; 52% (n=585) NHW, 22% (n=252) NHB, and 22% (n=246) Hispanic. The race/ethnic breakdown of the study population is similar to the distribution of race/ethnicity statistics, specifically that of NHW, NHB, and Hispanic within the state of Florida, and comparable to the NHW, NHB, and Hispanic distribution within the United States.^22^ The majority of study participants (61%, n=660) experienced a mild stroke (NIHSS 0-5). The frequency of moderate (NIHSS 6-15) and severe (NIHSS 16+) strokes was higher in NHB (26% moderate or severe) and Hispanic (26%) individuals, than for NHW (17%). Additionally, the study sample was primarily comprised of individuals who were able to walk independently at time of discharge (78%, n=849), and had either private insurance (23%, n=250) or Medicare (44%, n=482).

Of the 1,083 individuals included in the sample, 53% (n=576) received some form of rehabilitation services (inpatient and/or outpatient rehabilitation), with 5% (n=55) receiving only IRU, 33% (n=361) receiving only OP, and 15% (n=160) receiving both IRU and OP services. At 30-days after discharge, 70% (n=755) individuals demonstrated no functional change, 13% (n=145) improved, and 17% (n=183) worsened. At 90-days after stroke, 65% (n=699) of individuals showed no functional change from discharge, with 21% (n=227) showing improvements, and 15% (n=157) worsening. Details of participant characteristics are included in **Table 1**.

**Table 1:**
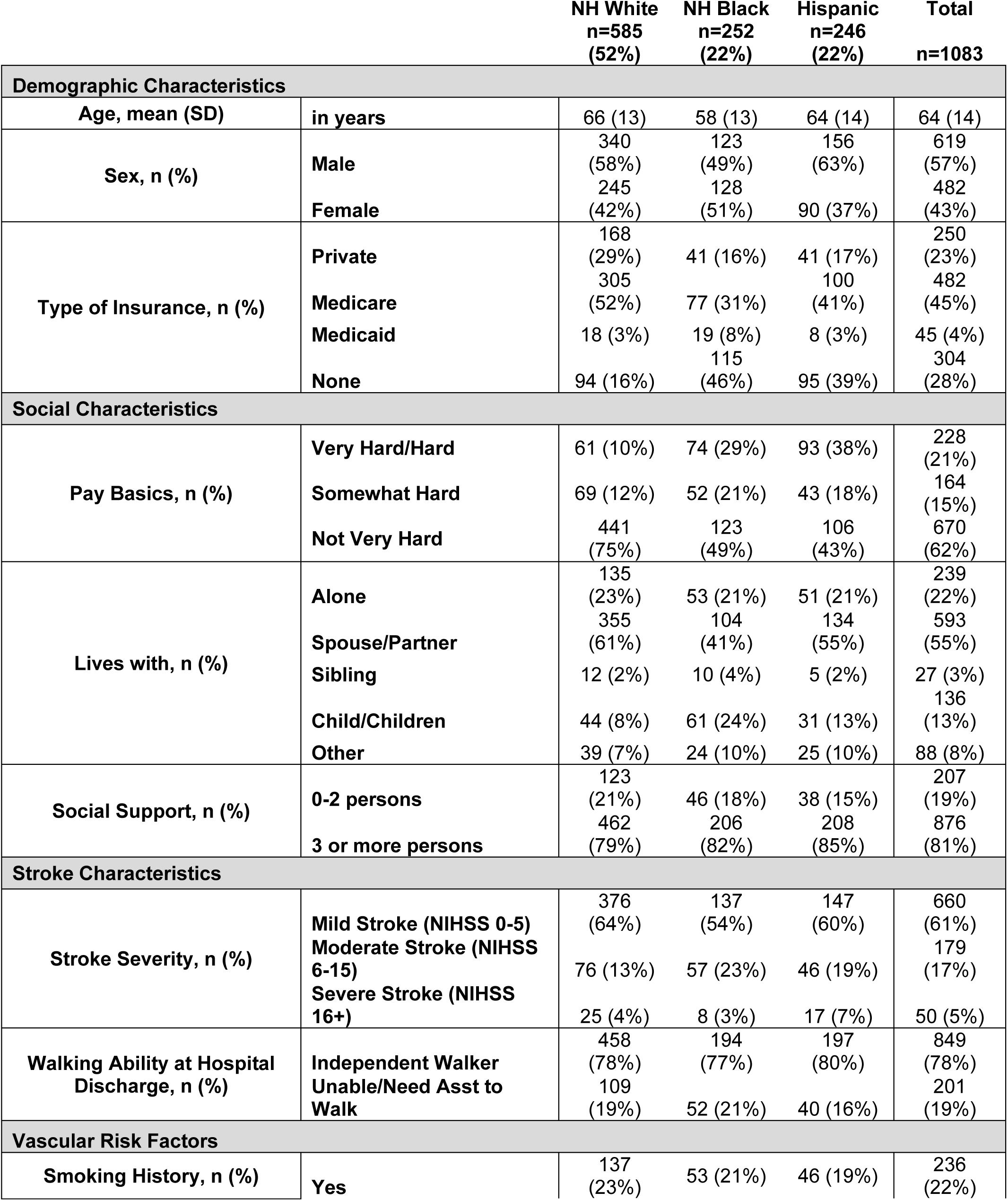
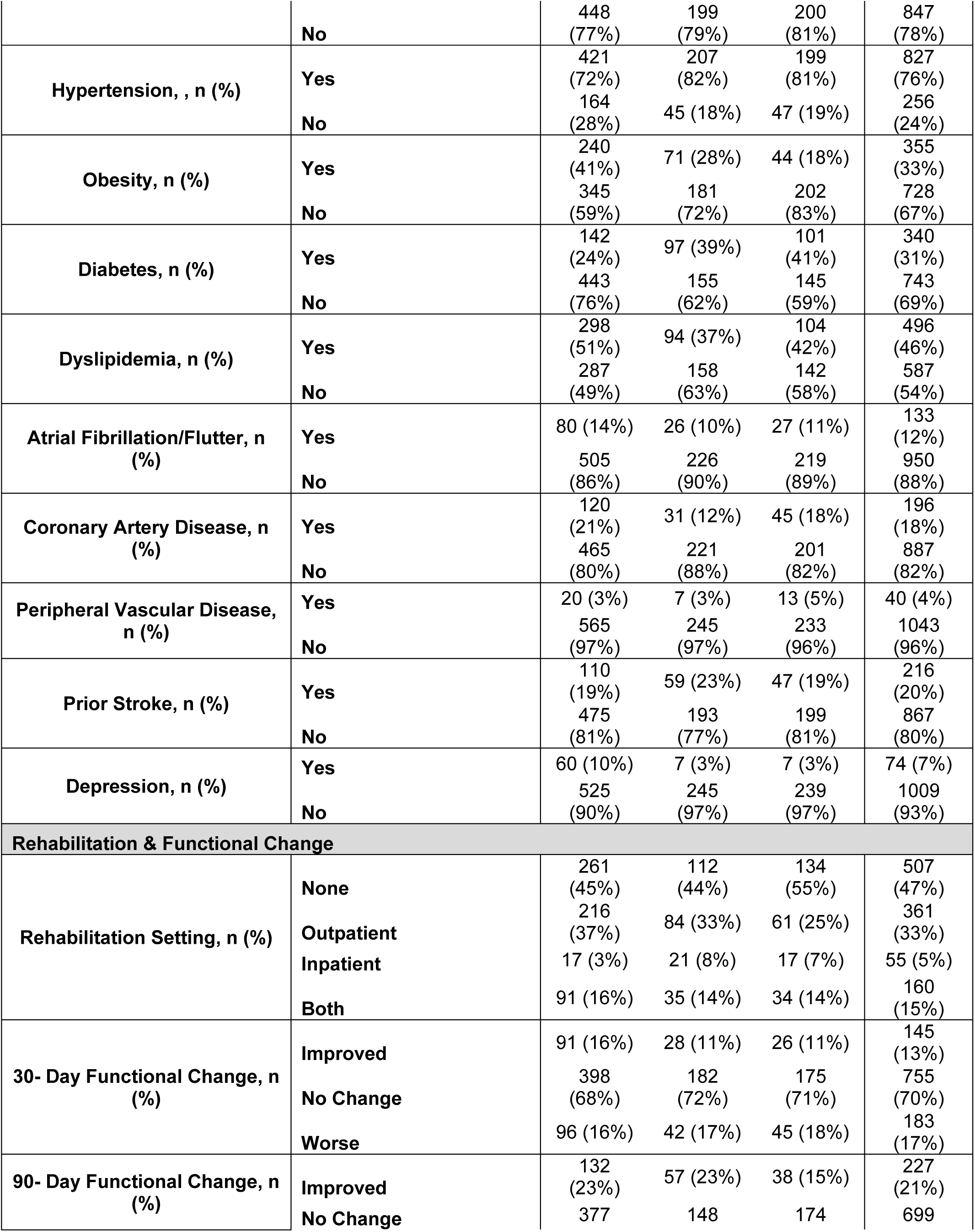
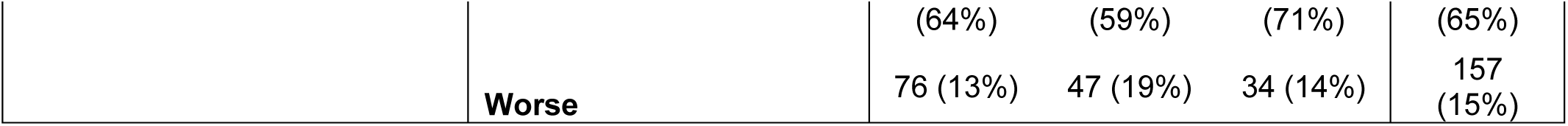
Participant Characteristics.

### The influence of race/ethnicity on rehabilitation services received

Hispanic individuals were significantly less likely to receive rehabilitation services (of any type) as compared to NHW [aOR=0.626, 95%CI (0.442,0.886)]. Additionally, Hispanic individuals were half as likely [aOR=0.543, 95%CI (0.368,0.800)] to receive OP services as compared to NHW. There were no other differences between NHW and Hispanic individuals. Further, there was no difference between NHW and NHB in respect to who received rehabilitation services and who did not, nor in respect to type of rehabilitation services received. **Figures 2A** and **2B** illustrate whether or not rehabilitation services were received (**2A**) and the type of rehabilitation services received (**2B**) by race/ethnic group. Results of the statistical models have been collated and are included in **Table 2**.

**Figures 2A and B:**
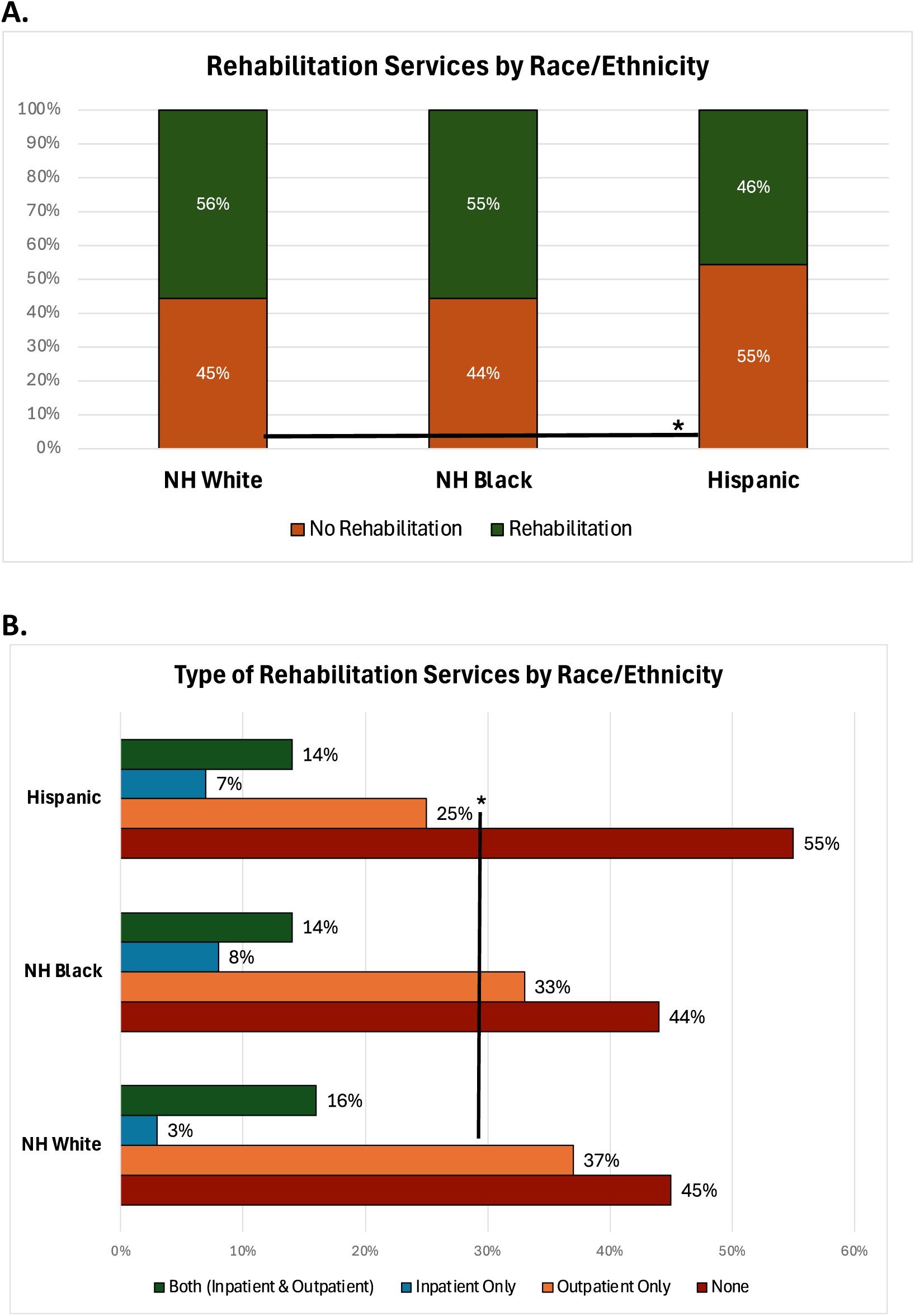
Race/ethnic differences in (A) receipt of rehabilitation services or not receiving any rehabilitation services and (B) type of rehabilitation services received.

**Table 2:** Associations between Receipt of Rehabilitation Services (any type of rehabilitation service vs none; type of rehabilitation services received) and Race/Ethnicity after hospital discharge.

**2. Rehabilitation Services By Race/Ethnicity**
| **REF is No Rehab** | **Effect** | **95% CI** |
| --- | --- | --- |
| Any Rehab - White | REF | REF |
| Any Rehab - Black | 0.996 | (0.699-1.421) |
| **Any Rehab - Hispanic** | **0.626** | **(0.442-0.886)\*** |
| Both - White | REF | REF |
| Both - Black | 0.919 | (0.521-1.619) |
| Both - Hispanic | 0.752 | (0.434-1.301) |
| IRU - White | REF | REF |
| IRU - Black | 1.971 | (0.885-4.388) |
| IRU - Hispanic | 1.294 | (0.579-2.892) |
| OP - White | REF | REF |
| OP - Black | 0.951 | (0.648-1.397) |
| **OP - Hispanic** | **0.543** | **(0.368-0.800)\*** |

\* Denotes statistical significance
†Model adjusted for: age, sex, insurance, stroke severity (NIHSS), walking ability at discharge, social network size, who patient lives with, ability to pay for life basics, history of prior stroke.
‡ Abbreviations: 'Any Rehab' refers to receipt of any configuration (Both, IRU, or OP) of rehabilitation services. 'Both' refers to receipt of both inpatient and outpatient rehabilitation services; 'IRU' represents receipt of inpatient rehabilitation services only; 'OP' represents receipt of only outpatient rehabilitation services.

### The influence of race/ethnicity on functional recovery

At 30-days post-stroke, Hispanic individuals were less likely to improve as compared to NHW [aOR=0.609, 95%CI (0.373,0.993)], with no statistically significant difference noted between NHB and NHW. Similarly, at 90-days after stroke, Hispanic individuals were less likely to show functional improvements [aOR =0.647, 95%CI (0.425,0.983)] compared to NHW. There was no statistically significant difference between NHB and NHW. There were no differences between race/ethnic groups in terms of worsening compared to no change from discharge to 30-days nor 90-days after stroke. **Figures 3A** and **3B** highlight race/ethnic differences in functional recovery at 30- and 90-days, respectively, after stroke. **Table 3A** contains results of statistical modeling.

**Figures 3A and 3B:**
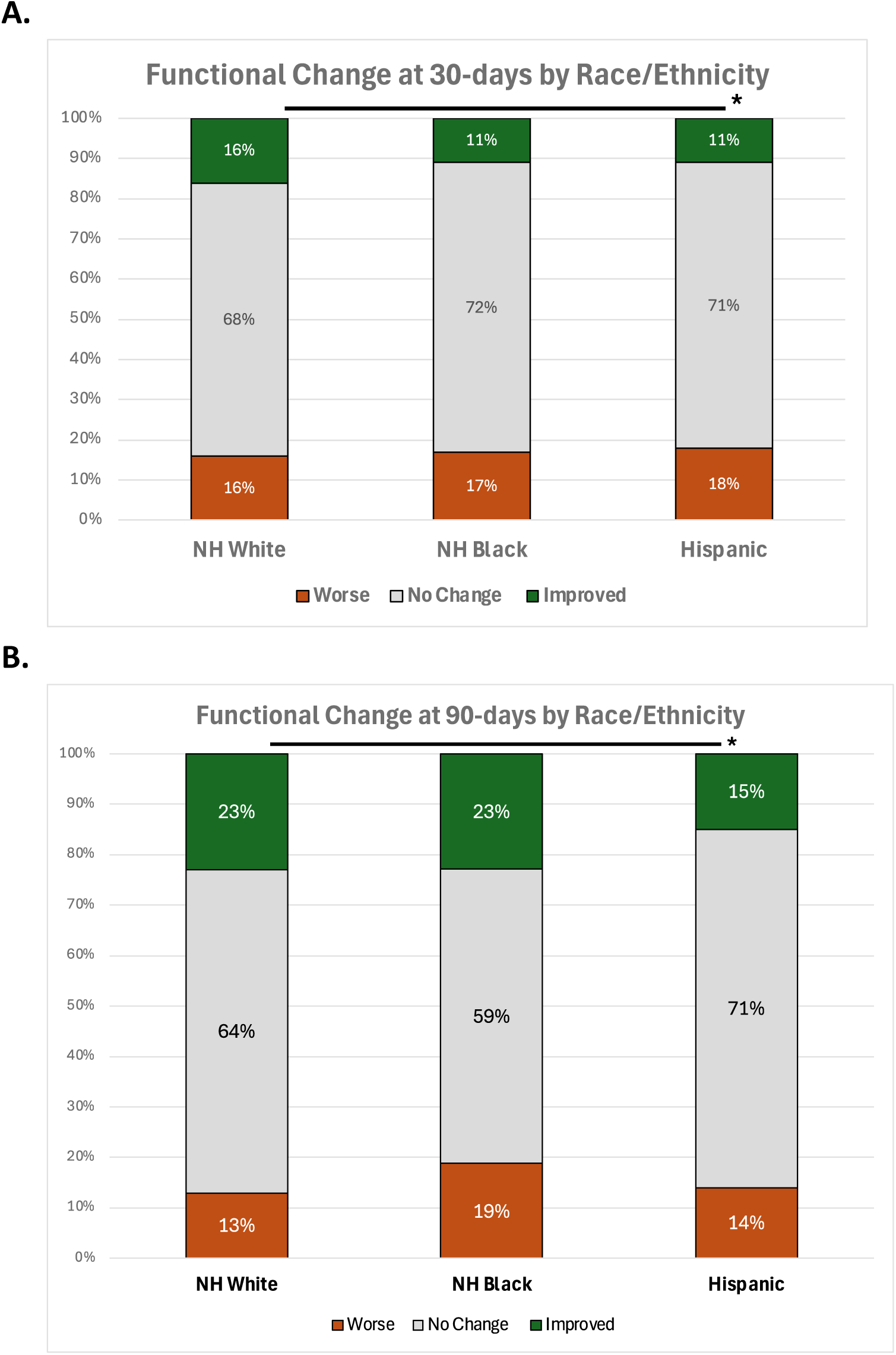
Race/ethnic differences in functional change at A.30- and B. 90- days after hospital discharge.

**Tables 3A and 3B:** Association between A.Race/Ethnicity and Rehabilitation Services Received (any type of rehabilitation service received vs none) with Functional Change at 30- and 90-days after hospital discharge.

**3A. Functional Change By Race/Ethnicity**
| 30-days | Effect | 95% CI |
| --- | --- | --- |
| Improved - White | REF | REF |
| Improved - Black | 0.701 | (0.428-1.149) |
| **Improved -Hisp** | **0.609** | **(0.373-0.993)\*** |
| Worsened - White | REF | REF |
| Worsened - Black | 0.821 | (0.527-1.278) |
| Worsened - Hisp | 0.960 | (0.627-1.468) |
| **90-days** |  |  |
| Improved - White | REF | REF |
| Improved - Black | 1.120 | (0.754-1.663) |
| **Improved -Hisp** | **0.647** | **(0.425-0.983)\*** |
| Worsened - White | REF | REF |
| Worsened - Black | 1.538 | (0.975-2.426) |
| Worsened - Hisp | 0.861 | (0.537-1.379) |

**3B. Functional Change By Rehabilitation Services**
| 30-days | Effect | 95% CI |
| --- | --- | --- |
| Improved | 0.995 | (0.675-1.467) |
| **Worsened** | **1.635** | **(1.151-2.323)\*** |
| **90-days** |  |  |
| **Improved** | **1.820** | **(1.301-2.545)\*** |
| **Worsened** | **1.631** | **(1.122-2.372)\*** |

REF is 'No Change' in function
\*Denotes statistical significance
†Propensity Score includes: Personal factors (age, sex, race, insurance), Social Factors (social support, lives with, pay basics), Clinical Factors (NIHSS, walk ability at hospital dc), and Vasclar Factors (DM, HTN, CAD, PVD, smoker, a fib/flutter, dyslipidemia, depression, prior stroke)

### The influence of rehabilitation on functional recovery

At 30-day follow up, individuals who went to rehabilitation were more likely to worsen [aOR=1.635, 95%CI (1.151,2.323)] as compared to those who did not receive rehabilitation services, regardless of services type. There was no difference in improvement between those who did and those who did not attend rehabilitation at 30-days. At 90-days after hospital discharge, individuals who attended rehabilitation were almost twice as likely to improve [aOR=1.820, 95%CI (1.301,2.545)], yet those who attended rehabilitation were also more likely to worsen [aOR=1.631, 95%CI (1.122,2.372)]. **Figures 4A** and **B** illustrate differences in functional recovery at 30- and 90-day follow up, respectively, between individuals who received rehabilitation services and those who did not. Results of statistical modeling are included in **Table 3B**.

**Figures 4A and 4B:**
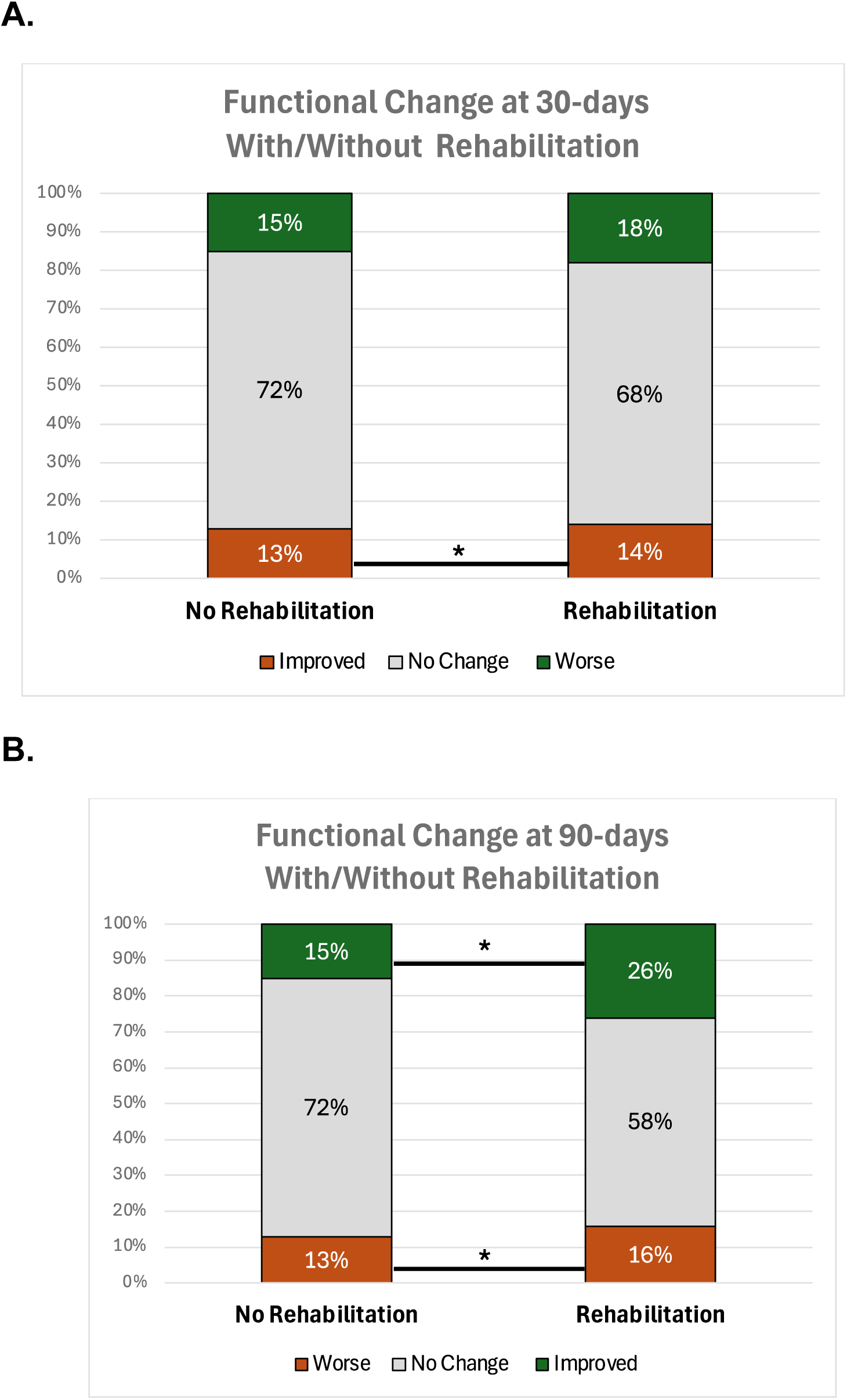
The association between rehabilitation services and functional change at A.30- and B. 90-days after hospital discharge.

## Discussion

The primary purpose of the TSCD-S was to investigate disparity in transitions of stroke care once individuals were discharged from hospital care as these patients are often ‘lost’ in the care system and highly affected by SDH. Rehabilitation aims to remediate motor impairments, promote physical activity (PA), and restore home and community function, thus reducing the risk of future stroke.^1,3,9,10,12^ However, only approximately half of our sample received rehabilitation services after discharge, with fewer Hispanic individuals receiving rehabilitation services.

Race/ethnic disparities in post-stroke care can impede access to rehabilitation services, the type of rehabilitation services received, and ultimately negatively impact outcomes in respect to return-to-home and community function. Our results have reiterated the benefits of rehabilitation on functional recovery after stroke and have further highlighted race/ethnic disparities in care access and type of rehabilitation services, particularly for Hispanic stroke survivors.

Consistent with prior work^23,24^, results of our study indicate that at 90-days after stroke, individuals who received rehabilitation were more likely to show improvements as compared to those who did not receive rehabilitation, irrespective of race/ethnic group. The use of a propensity score limited potential confounding by indication, though residual confounding remains a potential source of bias. Historically, race/ethnic disparities in functional outcomes have been noted between NHW and NHB stroke survivors.^19,25,26^ Yet, our findings did not show differences between NHW and NHB. There were no significant differences between NHW and NHB in functional improvements from discharge to 30-days and from discharge to 90-days after stroke. Given the increased incidence and increased stroke severity previously documented in NHB individuals, this gives us promise that we are moving toward health equity in stroke care.^16,17^ With respect to Hispanic stroke survivors, earlier work has noted race/ethnic disparities specifically in functional outcomes.^27,28^ Our results echo these findings. Compared to NHW, our results indicate that Hispanic individuals were only half as likely to demonstrate mRS improvements from discharge to 30-days and from discharge to 90-days. Previous work has cited the contribution of stroke severity and additional sociodemographic factors to stroke outcomes.^17,25,28–30^ However, we controlled for stroke severity, age, sex, insurance type, and receipt of rehabilitation services, as well as social network size, ability to pay for basic needs, and who the patient lives within our statistical modeling. While we have made strides to close the gap between NHW and NHB individuals, Hispanic individuals continue to remain at a disadvantage in respect to functional recovery.

We found no differences between NHW and NHB individuals in those who did/did not receive rehabilitation. Hispanic individuals, however, were less likely to receive any type of rehabilitation service after stroke, echoing the underutilization of rehabilitation services that was highlighted from work of more than 30 years ago.^31^ In a further breakdown of type of rehabilitation services received, no statistically significant differences were noted between NHW and NHB individuals. Given the prior influence of stroke incidence, stroke severity and sociodemographic factors^15–19,26,29–31^, our statistical modeling controlled for potential confounders of age, sex, insurance, stroke severity, social support, ability to pay for basic life necessities, and history of prior stroke. However, even after adjusting for these potential confounders, Hispanic individuals were half as likely to receive outpatient rehabilitation services compared to NHW. Our results suggest that while we are making strides towards improving health equity, we have not fully mitigated the gap between post-stroke services provided to NHW and services provided to people of color, particularly for individuals of Hispanic ethnicity.

In addition to the main effects reported in our results, we further evaluated for an effect modification. There was no significant interaction between race/ethnicity and rehabilitation on functional change, neither at 30-days nor at 90-days after stroke. Due to limitations in overall sample size, we further performed a stratified analysis to examine the influence of rehabilitation on functional outcomes by race/ethnicity. Results of the stratified statistical modeling are included in **Supplemental Table 1A** (NHW), **1B** (NHB), **1C** (Hispanic). From our stratified analysis, we can further interpret the results of our main effects, namely that the main effects were driven primarily by the NHW individuals included in this cohort as >50% of our cohort was NHW. Future studies should be conducted in large, multi-ethnic populations to further investigate the potential for effect modification by race/ethnicity for the association between rehabilitation utilization and functional change after stroke.

### Strengths and Limitations

Our study has several notable strengths. Our study captured a diverse race/ethnic group. Further, recruitment from participating hospitals throughout the state in both rural and urban areas yielded a diverse study population, improving generalizability beyond a single institution. An important limitation to the generalizability of our study results is that the majority (61%) of our participants experienced a mild stroke. In an effort to control for the mismatch of stroke severity between NHW, NHB, and Hispanic individuals, statistical modeling did adjust for stroke severity. Yet, it is likely that individuals with moderate or severe strokes would have different rehabilitation needs and may require more than 90-days post-stroke to demonstrate functional improvement. Regardless, there were no differences in stroke severity between NHB and Hispanic individuals, and recovery outcomes of NHB were similar to outcomes of NHW.

Another limitation to the study is that there was no record of the prescription of inpatient rehabilitation services, nor information on the prescription or use of home health rehabilitation services. It is plausible that inpatient rehabilitation may have been prescribed and not utilized or that other types of rehabilitation services may have been preferred, and our results may not reflect race/ethnic disparities but reflect patient preferences for rehabilitation services that were not fully measured. Therefore, we do not have the full picture of rehabilitation prescription and utilization. Other limitations of this study include the residual confounding by indication, and the additional confounding socioeconomic factors not included in this study. Further, the use of the mRS as a measure of functional recovery also has limitations. The mRS is a highly reliable scale that is frequently used as a marker of disability and/or function post-stroke.^28,32–36^ However, the mRS is a gross scale and as a result, the interpretation or the generalizability of these study results, specifically as related to mobility function is limited. Future work should include a marker specific to locomotor independence, such as the Functional Ambulation Category^37^, in order to more accurately identify characteristics that contribute specifically to locomotor recovery and explain home and community-level PA, specifically with an eye to improve activity levels and reduce risk of recurrent stroke.^1–3^ Future work should additionally consider methods that directly observe home and community-level locomotor behaviors, such as smartphone applications or IMUs, that could extend both evaluation and even potentially rehabilitation services to mitigate disparities in underserved race/ethnic groups who may have limited resources or access to rehabilitation.

### Conclusions

In summary, our results emphasize the important role rehabilitation plays in functional recovery after stroke. Yet, race/ethnic disparities remain. We have made strides to mitigate the equity gap between the functional outcomes of NHW and NHB individuals. However, Hispanic individuals were half as likely to demonstrate functional gains 30-days after hospital discharge. This disparity in functional improvements was maintained at 90-days. Further, race/ethnic differences persist between types of rehabilitation services utilized. Hispanic individuals remain underserved and are less likely to receive outpatient services after discharge.

## Data Availability

Data included were a subset of a larger stroke registry (Florida Stroke Registry). Data availability will be considered on request.

